# Scoping review of online postal sexually transmitted infection services: access, usage and clinical outcomes

**DOI:** 10.1101/2021.12.10.21267608

**Authors:** Kirsi Sumray, Karen C. Lloyd, Claudia Estcourt, Fiona Burns, Jo Gibbs

## Abstract

**Background:** There has been considerable expansion in online postal self-sampling STI services in many parts of the UK, driven by increasing demand on sexual health services and developments in diagnostics and digital health provision. This shift in service delivery has occurred against a backdrop of reduced funding and service fragmentation and the impact is unknown. We explored characteristics of people accessing and using online postal self-sampling (OPSS) services for STIs in the UK, the acceptability of these services, and their impact on sexual health inequalities.

**Methods:** A scoping review was conducted of studies published in English-language based on pre-agreed inclusion/exclusion criteria, between 01/01/2010 to 07/07/2021. Nine databases were searched, and 23 studies that met the eligibility criteria were included. Studies were appraised using the Mixed Methods Appraisal Tool.

**Results:** Study designs were heterogenous, including quantitative, qualitative and mixed method analyses. The majority were either evaluating a single site/self-sampling provider, exploratory or observational and of variable quality. Few studies collected comprehensive user demographic data. Individuals accessing OPSS tended to be asymptomatic, of White ethnicity, women, over 20 years, and from less deprived areas. OPSS tended to increase overall STI testing demand and access, although return rates for blood samples was low, as was test positivity. There were varied results on whether services reduced time to treatment. OPSS services were acceptable to the majority of users. Qualitative studies showed the importance of trust, confidentiality, discretion, reliability, convenience and improved patient choice.

**Conclusion:** OPSS services appear highly acceptable to users. However, uptake appears to be socially patterned and some groups that bear a disproportionate burden of poor sexual health in the UK are under-represented among users. Current provision of online self-sampling could widen health inequalities, particularly where other options for testing are limited. Work is needed to fully evaluate the impact and cost-effectiveness of OPSS services.

**Key messages**

1. An increasing proportion of STI testing in the UK is occurring via online postal self-sampling services
2. Service users tend to be asymptomatic, white, women, over 20s, and from less deprived areas
3. OPSS services are acceptable to users and can improve choice
4. There is a need for a wide-ranging evaluation of OPSS services to determine their impact on sexual health inequalities, access, clinical outcomes and service delivery

## INTRODUCTION

“Home testing” whereby an individual tests for sexually transmitted infections (STIs) and/or blood borne viruses (BBV) remote from traditional healthcare settings, encompasses user self-sampling (the laboratory processes and tests the samples) or self-testing (the user tests their own samples) (1). The most widely used option in the UK is online postal self-sampling (OPSS) in which users order specimen collection kits via the internet. Kits are delivered by post or may be collected from a sexual health clinic. Users obtain their own samples (typically urine, vulvo-vaginal, ano-rectal and pharyngeal swabs), and repackage before posting back to a laboratory for testing. Test results are made available by text message or online. OPSS is increasingly provided in high income countries (2–4). England’s earliest online chlamydia testing services began in 2006 (5), but the other devolved nations of the United Kingdom (UK) introduced OPSS rather later (6–8). The British Association for Sexual Health and HIV (BASHH) recommends online testing and care provision as an adjunct to in-person services to increase choice for service users (9).

The UK had seen a sustained rise in several STIs until the start of the COVID-19 pandemic (10), and demand for sexual health service (SHS) consultations has increased. Between 2015 and 2019 in England there was a 23% increase (3,143,144-3,852,121). Total sexual health screens for chlamydia, gonorrhoea, syphilis and HIV increased by 31% (1,657,425 to 2,175,525) in this period. Internet based services have assumed an increasing proportion of consultations and screening activity. In 2018-19, while overall consultations and tests in England rose by 7% and 10% respectively, consultations and STI screens provided by internet-based services surged by 94% and 69% (11). This trend of an increasing proportion of testing and consultations being accessed and provided online has accelerated as a result of the COVID-19 pandemic (10). Young people, people from ethnic minority groups, men who have sex with men (MSM), people who are gender diverse, and those living in more deprived areas have borne a disproportionate burden of STIs and poorer sexual health for many years (12–17). The reasons for this disparity are not sufficiently understood (18).

Some evidence suggests that OPSS might improve access to STI services for those who feel uncomfortable or struggle to attend face-to-face services (19), and might cost less than in-person care (20,21). However, relatively little is known about the characteristics of people who use OPSS and the existing literature is heterogeneous and has not previously been comprehensively reviewed. With a drive towards further online sexual health provision in the UK (22), the pre-existing unequal burden of sexual ill health in the population, and well described inequalities in access to sexual health care, it is important to understand the impact of this shift in service delivery. We aimed to evaluate the current evidence on access to and usage of OPSS services. We have restricted our scope to the UK because online care has been relatively mainstreamed in at least one country (England) for many years and sexual health care is provided free at the point of access without the need for specialist referral. Our specific objectives were to: 1. describe characteristics of people accessing and using STI self-sampling services in the UK; 2. assess whether OPSS increases demand for testing; 3. assess the impact of OPSS on clinical outcomes; 4. assess levels of acceptability of OPSS services in the UK.

## METHODS

We conducted a systematic scoping review in order to map and synthesise the current research evidence, in an area where the existing literature is heterogeneous and has not previously been comprehensively reviewed (23). We followed the Preferred Reporting Items for Systematic Review and Meta-Analyses extension for scoping reviews (PRISMA-SCR) guidelines (24). A review protocol has not been registered. The PICO framework was used to identify the research question and objectives:

### Population

People residing in the UK who are engaging in sexual activity and accessing online sexual health services.

### Intervention

Self-sampling STI testing kits which are posted to individuals’ homes by online sexual health services.

### Comparators

The alternatives to this intervention are for individuals to visit sexual health clinics, General Practice, or community outreach services to be tested by healthcare professionals, self-sample in a clinical setting, or collect kits to bring home. This review will compare OPSS with these comparators where included studies have done so.

### Outcome

Accessibility and usage of OPSS services with a focus on inequalities in access and acceptability of services.

### Study selection criteria

Studies were included if they described the use of home self-sampling kits provided by online sexual health services in the United Kingdom, published between 1.1.2010-30.06.2021 in the English language. All non-theoretical study types with full-text online access were included. Studies were excluded if they described services outside of the UK, published prior to 2006 or not in English, and not focusing on home self-sampling kits provided by online sexual health services (for example, STI education, contraception or self-testing kits). We also excluded “siloed” (HIV only) HIV self-sampling services because the national HIV self-sampling service targets specific key populations such as MSM and black African groups (22) rather than the general population, and one of our key objectives was to evaluate access to, including potential inequalities with, the use of OPSS services. Protocols of studies and research displayed via conference or other forms of presentation were excluded if there was no full-text access.

### Study selection

De-duplication and title screening was conducted by the first author; 20% of the abstract screening and 100% of the full-text screening were verified by a second reviewer (KCL and JG, respectively) independently to reduce selection bias (25). The inter-reliability rate of the two reviewers was over 90% at the abstract-screening stage and was 100% for full-text screening.

### Search method

A systematic search and data extraction was conducted on 22^nd^ June 2020 to fulfil requirements of KS’ Masters dissertation and was rerun on 7^th^ July 2021 by JG in nine databases: Medline, EMBASE, PsycInfo, The Health Management Information Consortium (HMIC), Web of Science, CINHAL Plus, Scopus, Open Grey and Ethos. The use of two grey literature databases (Open Grey and Ethos) aimed to reduce potential publication bias and provide a more comprehensive view of the evidence (26). The search consisted of a selection of medical subject headings (MeSH terms), where appropriate, and free-text. Limits were used in applicable databases which limited by date and language. The search comprised of four concepts: types of STI, type of online or self-sampling service, accessibility or inequalities, and UK filters. For example, terms such as ‘STI’, ‘Chlamydia’, ‘Self-Sampl*’, ‘eHealth’, ‘Access*’, ‘Inequalit*’, ‘United Kingdom’, and ‘England’ were used. To ensure the search strategy was fully comprehensive, additional terms for ‘eHealth’ were included (27) and adapted and simplified versions of two verified filters for the UK were used in database searches, where appropriate (28,29).

The search only included studies published between 2010-2021; this was due to the very low numbers of users of OPSS services prior to 2010 (5). See online supplemental file 1 for details of the full search strategy.

### Data extraction

The phases of data identification, de-duplication, screening and eligibility checks shown in Figure 1. For included studies, a data extraction form (online supplemental file 2) was used to extract the demographics of the study participants (online supplemental file 3), the study design and key findings from each study (online supplemental file 4).

**Figure 1:**
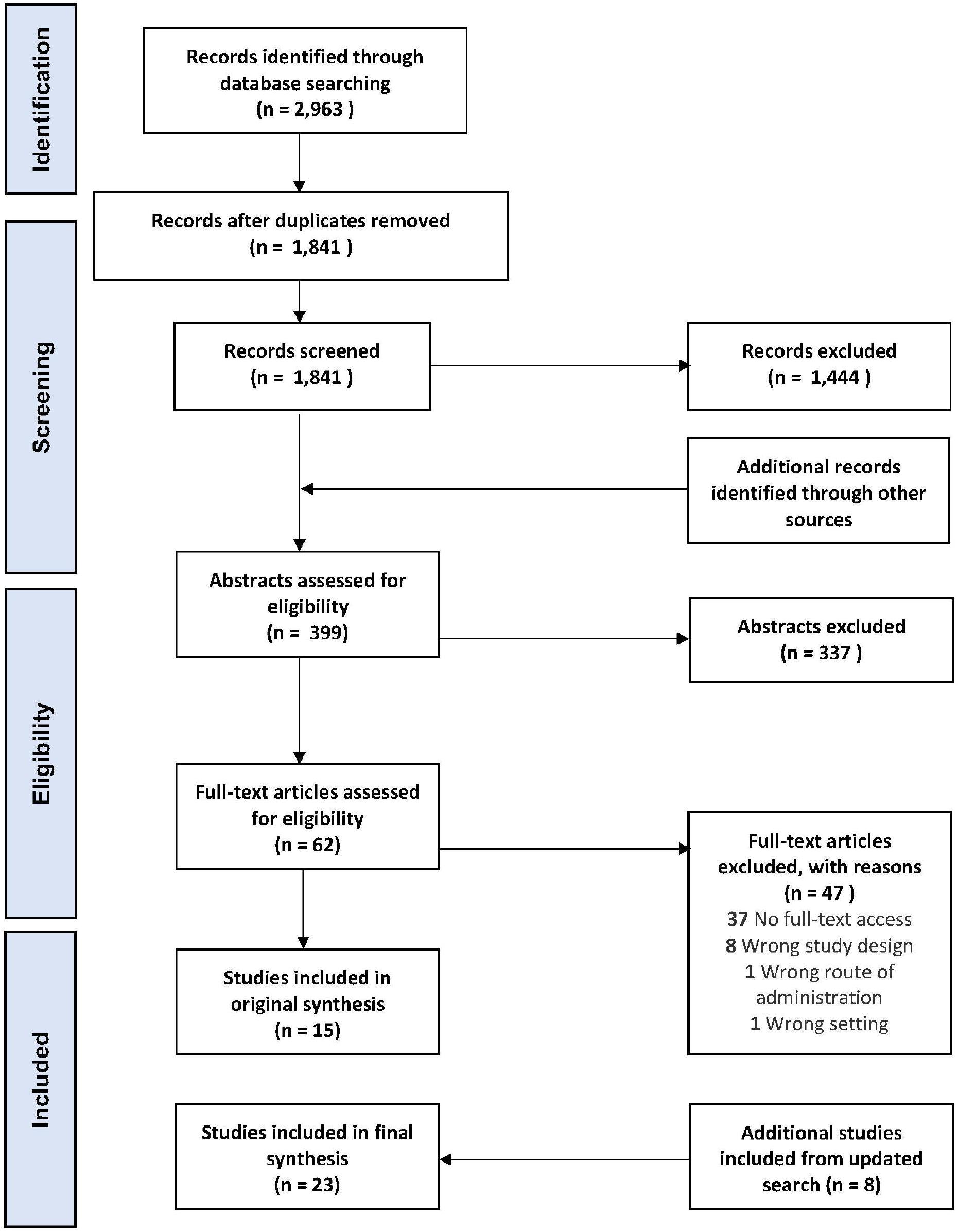
Flow of Information, based on the PRISMA Flow Diagram (32)

### Empirical appraisal and analysis of included articles

Study designs were heterogeneous, including quantitative, qualitative and mixed method analysis, and were therefore appraised using the Mixed Methods Appraisal Tool (MMAT) (30). Quantitative data were analysed using descriptive statistics, and qualitative data were analysed using inductive thematic analysis (31).

## RESULTS

### Overview of included studies

This search strategy identified 23 relevant articles that described ten different OPSS services, all located in England (see Table 1 and online supplemental file 3 and 4). The overall quality was variable, with the majority either evaluating a single site/testing provider, and exploratory or observational. See online supplemental file 5 for the full MMAT results. For a summary of each individual service or intervention, see online supplemental file 6. Six studies explored solely chlamydia testing, one assessed a chlamydia and gonorrhoea testing service and seventeen assessed services which tested for chlamydia, gonorrhoea, HIV and syphilis. Five articles compared demographic characteristics and outcomes of users of OPSS with clinic-based services (20,32–36).

**Table 1:** Overview of Included Studies.

| Service | Author (Year) | Study Type | Study Aims | Demographic characteristics captured |  |  |  |  |
| --- | --- | --- | --- | --- | --- | --- | --- | --- |
|  |  |  |  | Gender | Age | Ethnicity | Sexual orientation | IMD |
| eSexual Health Clinic (eSHC) | Aicken et al. 2018 (37) | Qualitative interviews | To understand use and experience of the eSHC to inform future evaluation and refinement | Binary | 18-35 | ✓ | ✓ | - |
|  | Estcourt et al. 2017 (38) | Non-randomised, exploratory proof of concept study | To assess the safety and feasibility of eSHC | Binary | 18+ | ✓ | ✓ | - |
|  | Gibbs et al. 2018 (39) | Mixed methods evaluation | To evaluate the eSHC results service | - | - | - | - | - |
| Freetest.me | Dolan et al. 2014 (40) | Randomised experiment | To explore the effect on chlamydia test return rates of non-cash financial incentives, and the influence of socioeconomic status | Binary | 16-24 | ✓ | - | Mean score |
| Letstalkaboutit | Gasmelsid et al. 2021 (34) | Retrospective service evaluation | To determine whether online screening is accessible by those patients most at need by comparing the demographics and number of asymptomatic chlamydial infections detected online and in clinic. | Binary | <25, 25+ | ✓ | ✓ | -** |
| NCSP | Woodhall et al. 2012 (5) | Quantitative retrospective data analysis; website evaluation | To describe and evaluate access to the NCSP's online chlamydia testing service | Binary | 15-24 | ✓ | - | ✓ |
| North East Essex PCT | Bracebridge et al. 2012 (41) | Cross-sectional study | To quantify uptake and test-positivity rates, identify factors associated with screening and compare costs of the intervention with the NCSP | Binary | 17-25 | ✓ | - | ✓<sup>#</sup> |
| Saving Lives | Page et al. 2019 (42) | Observational study | To ascertain how DBS HIV kits compared with MT kits in this postal testing service. | Female, male, trans | Median 26 | ✓ | ✓ | - |
| Sexual Health London | Day et al. 2020 (43) | Retrospective service evaluation | To report the rate of recent sexual assault (SA) disclosure amongst users of SHL, and identify the outcomes of their call back discussions. | Female, male, trans or non-binary | 18-55 | ✓ | ✓ | - |
|  | Day et al. 2021 (44) | Retrospective service evaluation | To assess the sexual health needs, sexual practices, STI/HIV positivity and satisfaction rates of trans and non-binary users of Sexual Health London | Female, male, trans, non-binary/gender fluid | 15-82 | ✓ | - | - |
|  | Day et al. 2020 (45) | Retrospective service evaluation | To report the safeguarding concerns and outcomes of 16-17year olds accessing SHL | Female, male, trans or non-binary | 16-17 | ✓ | ✓ | - |
|  | Day et al. 2021 | Retrospective service | To identify the characteristics and transfer to care | Binary | 21-50 | ✓ | ✓ | - |

|  | (46) | evaluation | rates of those who have a reactive HIV test result via SHL |  |  |  |  |  |
| --- | --- | --- | --- | --- | --- | --- | --- | --- |
| SH:24 | Barnard et al. 2018 (32) | Cross-sectional study | To compare the characteristics of e-STI service users with clinic users, and OPSS kit returners with non-returners | Binary | 16+ | ✓ | ✓ | ✓ |
|  | Barnard 2020 (Chapter 6) (47) | Qualitative interviews | To describe the experiences, barriers and facilitators of SH:24 in Lambeth and Southwark | Female, male, trans | 16-30 | ✓ | ✓ | - |
|  | Syred et al. 2019 (48) | Observational study | To describe user choice of OPSS orders and diagnoses in a 'choose to test' intervention | Binary | 16-24 | ✓ | ✓ | ✓ |
|  | Turner et al. 2018 (20) | Observational study | To investigate the effect of decision-making on resource allocation in a clinic after the introduction of an e-STI service in Lambeth and Southwark | Binary | 16+ | ✓ | ✓ | - |
|  | Turner et al. 2019 (33) | Case study analysis and model generation | To establish cost-effectiveness of an OPSS service, and explore cost per diagnosis in different scenarios | - | - | - | - | - |
|  | Wilson et al. 2017 (49) | Single-blind randomised control trial (RCT) | To assess the effectiveness of an OPSS service compared with face-to-face services | Female, male, trans | 16-30 | ✓ | ✓ | - |
|  | Wilson et al. 2019 (50) | Secondary data analysis of an e-STI RCT in Lambeth and Southwark | To examine the effect of an e-STI service on testing uptake on people who had never previously tested (never-testers) | Female, male, trans | 16-30 | ✓ | ✓ | - |
| TakeATestUK.com | Page et al. 2021 (51) | Observational study | To ascertain how DBS HIV and syphilis kits compared with MT kits in this postal testing service | Female, male, trans | Mean 27 | ✓ | ✓ | - |
| Umbrella | Banerjee et al. 2018 (35) | Retrospective service evaluation | To evaluate the rates of uptake and return of OPSS kits and compare patient demographics and clinical outcomes in home and clinic testers | Female, male, transgender | 16+ | ✓ | ✓ | - |
|  | Banerjee et al. 2020 (36) | Retrospective service evaluation | To evaluate the uptake, return rate and new diagnosis rates of home-based testing in comparison with clinic-based testing for HIV, syphilis and hepatitis B | Female, male, transgender | 16+ | ✓ | ✓* | - |
|  | Manavi et al. 2017 (52) | Observational study | To establish which factors influence return of OPSS kits | Female, male, transgender | - | - | - | IMD rank |

### Characteristics of those accessing OPSS services

Comprehensive demographic data was not consistently collected across studies, in terms of both type of data collected and what was collected (see Table 1). Only two studies collected demographic data on all of gender, age, ethnicity, sexual orientation and the Index of Multiple Deprivation (IMD) (32,48); two collected none of these items (20,39). In addition to age and gender, six studies collected IMD data, 17 collected sexual orientation and 21 collected ethnicity, the majority of which were described in different ways using a variety of groupings. Gender types captured also varied between studies, with two that did not collect this data, and ten only reporting binary types.

Those who accessed services tended to be majority women (56.7-69.4% women, five studies reported statistical significance) (5,20,32,34–36,40,41,51), were resident in less deprived areas compared to accessing testing in other settings (p<0.001) (5,32,40,41), white or white British (53.3-92% users, four studies reported statistical significance) (5,32,34–36,40,51), and 20 years old or over (95.4% when compared to any age group (20,32), 32-42.9% when c.f. people aged 20-25 (41,53)), with two studies reporting statistical significance (20,32).

In those studies that collected more comprehensive gender data, people who identified as gender diverse made up a small proportion of the overall population accessing the service (0.0-0.42%) (35,36,44,49–52).

There were limited and conflicting data on access according to sexual orientation; one study reported more MSM requesting access than heterosexual men (32), and one study found that a higher proportion of people identifying as non-heterosexual accessed online self-sampling compared to clinic based testing (OR 0.44, (95% CI 0.27-0.72) (34). This is in contrast to findings from the Umbrella service, Birmingham (36).

Those accessing online services sometimes displayed higher risk behaviours (5,40) but often had a greater proportion of negative test results (32,33,35,36). Individuals were more likely to order OPSS kits if they had used STI testing services before (40). Of those who were never-testers, a significant proportion were recruited face-to-face in communities (50).

### Usage of OPSS services

The proportion of users returning self-sampling test kits varied by study (range 48%-78.4%) (32,35,36,44). Women were more likely than men to access and return STI self-sampling kits in the majority of studies (20,32,35,36,40,41,52). One service evaluation found no difference between kit return rates between those identifying as gender diverse and those identifying as cisgender (OR 1.00, 95%CI 0.81-1.24), although successful return (OR 1.6, 95% CI 1.06-2.36) and successful testing (OR 1.2, 95% CI 0.87-1.66) of blood samples were higher in those identifying as trans or non-binary/gender fluid (44). Characteristics of those who returned test kits varied between studies for age (32,36,40), and by level of deprivation (40,52). However, across several studies, people of white ethnicity were more likely to return the kit compared to other ethnicities (32,35,36).

A study that evaluated those accessing an OPSS spontaneously, compared to those that attended clinic and were triaged to testing online, found that the return rate was slightly lower in the triage and signpost group compared to the spontaneous online group (67.0% vs 70.5% by 6 weeks respectively) (20).

There is limited data on return rate of blood sampling kits, with one study finding only 54% (9033/16,611) of people returned a blood sample with a sufficient quantity of blood for testing (36). Studies that examined different types of blood sampling kits found no difference between return rates for dried blood samples (DBS) versus mini tests (MT) (66.5% vs 68.7%) (42), but did find that the samples were significantly more likely to be successfully processed with DBS (94.6%-98.8%) compared with MT (55.7% -54.5%, p=<0.001) (42,51).

### Impact of OPSS services on demand

In many areas, the introduction of OPSS services was not associated with a change in numbers of people attending clinics, but increased the overall demand for STI testing services (20,33,49).

### Impact on clinical outcomes

#### Test positivity

Overall test positivity for chlamydia and gonorrhoea was lower in OPSS services (4.4-8%) than clinic-based services (10-14.4%) (32,35). Two studies found a low HIV prevalence in their OPSS testing population; 0.097% (144/148,257) (45) and 0.83% (75/16,611) (36) of users in the evaluation period had a reactive HIV result. Of these, 65% (36) and 92% (45) had confirmatory testing, and 1% (1/75) (36) and 23.6% (34/144) had a new diagnosis of HIV confirmed. When comparing DBS and MT for HIV self-sampling, MT was found to have a higher proportion of reactive tests (6.2% vs 0.5%) (51), lower proportion of confirmed reactive tests (n=1/30 (3.3%) vs (1/11) 9.1%) (51)and higher false positive tests (5.2-5.4% vs 0.0-0.4%) (42,51). However, these were service evaluations and the blood sampling kit options were offered sequentially rather than in parallel.

#### Time to treatment

There were varied results regarding time-to-treatment; one paper reported that online patients took longer to receive treatment than clinic users (35), and one study found that there was no statistically significant difference (49). When an entire care pathway was trialled online and asymptomatic chlamydia-positive individuals could access an automated online clinical consultation which allowed people to collect their treatment at a community pharmacy, median time to treatment was one day (IQR 0-1) (38). A study comparing outcomes of asymptomatic service users testing positive for chlamydia via clinic-based services and online found that those diagnosed online were less likely to wait more than a week for treatment compared to those diagnosed in clinic (OR 9.94, 95% CI 2.87-34.42) (34).

#### Reporting sexual assault and safeguarding outcomes

When evaluating outcomes of those people who reported online a recent sexual assault over a 6-month period in 2020, one service found that 0.5% (n=242/45841) of users indicated they had been a victim of a recent sexual assault, which led to telephone intervention by a clinical healthcare professional. Of these, nearly 80% of people were contacted. However, 41.7%, (n=101/242) of users stated that they had not intended to report recent sexual assault. Fifteen people had already reported the sexual assault and had been seen by the police or by a Sexual Assault Referral Centre (SARC). One person required a SARC referral, and 8 people were referred to a clinic-based service (43). The same service also evaluated their safeguarding outcomes in 16-17 year olds, finding that a high proportion of this population (42.5%) met the service criteria for a follow-up telephone call from a health advisor. The most frequent reason that a call was triggered was related to drug and alcohol use (27%). The outcome of 8.5% of calls was a discussion with the child protection team, with 7% requiring a referral or discussion with social services (45).

### Acceptability of OPSS services

Qualitative research showed the importance of trust, confidentiality, discretion, reliability, convenience and improved patient choice in ensuring the successful use of OPSS (37,47), and between 74-98% of individuals surveyed expressed that they were pleased with and found these services acceptable (39,49,50).

### Economics

Although this review did not specifically set out to evaluate the economic outcomes of OPSS services, only one study included any costings data (33). This study, evaluating the impact of online testing across specialist sexual health services in two London boroughs found that, although there was an increase in the total annual cost of STI testing following the introduction of online testing, the average cost per test and diagnosis decreased (33)

## DISCUSSION

Although evaluations of OPSS of variable quality were found, we did not identify any large scale, multicentre robust studies. Available evidence suggests that OPSS services appear to be most used by and acceptable to asymptomatic individuals who are predominately women, over 20 years of age, resident in less deprived areas, and of white ethnicities. There is preliminary evidence that people from groups experiencing a disproportionate burden of STIs use OPSS less than other groups. The heterogeneity of the included studies prevented full evaluation of clinical outcomes.

Online services tend to be targeted at asymptomatic individuals who do not have other sexual health needs. The ability for asymptomatic people to manage their care needs remotely is an important and useful contribution to detect symptomless STIs, in order to enable treatment of the index patient and partner notification, and reduce morbidity and onward transmission. However, asymptomatic users reported higher risk behaviours in some studies (5,40), and there was no data on the impact of using OPSS on future sexual behaviour. In addition, there was insufficient evidence to be able to establish the impact of OPSS on treatment and partner notification outcomes.

Women were more likely than men to access and return STI self-sampling kits in the majority of studies (20,32,35,40,41,52). For services that provided accessibility information by sexual orientation, MSM seemed to be successfully using OPSS services (32,52). This finding is consistent with those from an OPSS in Canada (54). Certain minority ethnic groups such as black Caribbean, black African and mixed ethnicities, who are also key populations that are at higher risk of poor sexual health were underrepresented in OPSS users (11), perhaps due to a preference for face-to-face care (47).

Some services appear to appropriately reach younger people, but other services were more popular with those aged over 20 (35). This could be because younger people are more likely to live at home and have concerns about parents finding a test kit delivered through the post (55). Using chlamydia as an exemplar, chlamydia has formed 49% of new STI diagnoses in 2019, yet there has been a 13% reduction in tests completed by young people since 2015 (11). Chlamydia is the most commonly reported STI in the UK, and disproportionately affects young people from deprived areas (53), so it is crucial to ensure services are targeting these groups within their region. Despite this, recent research shows that both men (adjusted odds ratio (aOR): 1.36 [95%CI: 1.35-1.39], p<0.001) and women (aOR: 1.32 [95%CI: 1.31-1.33], p<0.001) living in the least deprived quintile were more likely to use OPSS services for chlamydia screening than those from the most deprived quintile (56).

Understanding inequalities in access and usage of OPSS services requires services to collect comprehensive socio-demographic data. Of the fifteen included studies, only two collected gender, age, ethnicity, sexual orientation and IMD data (32,48). Most studies included categories such as ‘other’ ethnicity or sexual orientation and though a few included trans participants, only one described people of non-binary, gender fluid or other genders (44). This data gap results in not only a skewed understanding of the impact of STIs on minorities, but also “facilitates the erasure of communities” (18). Further, the interrelations between demographic groups have not been sufficiently examined in these studies despite reference in the literature (e.g. 4,11,65).

Return rates of blood samples that were sufficient for testing were relatively low, and in keeping with findings from the National HIV self-sampling service (58). Overall test positivity was lower in OPSS services compared to clinic-based services (32,36). This is in keeping with, but more marked than, findings from both selective and unselective national datasets (10,58). As highlighted within one study, the low PPV for home-based BBV testing is concerning (36) and requires further evaluation from both a cost-effectiveness and well-being perspective.

To our knowledge, this is the first review to explore the accessibility of OPSS services in the UK. The inclusion of qualitative as well as quantitative studies ensured that both acceptability and core themes regarding people’s access to and use of services could be explored. The included studies were heterogenous in design using a variety of methodologies which added value to the review. By excluding hypothetical studies, acceptability and barriers to service use are indicative of the real-life experience of online service users.

Fourteen of 23 studies took place in London and no studies were conducted in Wales, Scotland or Northern Ireland so findings may not be generalisable to all of the UK. This review did not set out to evaluate partner notification or economic outcomes of OPSS services, but the authors observed that there was a dearth of information on these within the studies that were included in this review. This study only focussed on the UK setting as it was focussing on access, and the infrastructure of sexual health service provision in the UK is different to other settings. However, the findings relating to acceptability and convenience are similar to those reported in systematic review and synthesis of qualitative research on OPSS services (55).

OPSS services appear acceptable to current users and improve choice but evaluation is limited. The existing evidence suggests that successful services achieve reliability, privacy, convenience, trust, and are integrated with clinic-based services to provide ease of transition between modalities of care. Online services are successful in relieving pressure from clinics (59), but clinic-based services remain essential for symptomatic individuals, people who have digital constraints, and those who prefer face-to-face care and or require additional safeguarding, amongst others (18).

With the extensive expansion in provision of OPSS services in recent years, a large-scale, multi-centre evaluation is needed to determine their cost-effectiveness and impact on access, clinical outcomes and service delivery. Clinical outcomes are key to understanding the cost-effectiveness of these services and there is limited research into this important factor. Further research is required to understand why people aged under 20 have lower uptake, and whether this is related to, for example, an individual’s experience of autonomy, competence and relatedness (60) and how awareness and access can be improved for these individuals (61).

## Supporting information

PRISMA checklist

## Data Availability

All data produced in the present study are available upon reasonable request to the authors

## The author declares no competing interests

FB, CSE and JG report receiving NIHR funding to research digital sexual health (NIHR129157 (FB, JG) and NIHR200856 (CSE, JG). CSE and JG are associate editors for STI journal.

## Contributions statement

KS conducted the original scoping review to fulfil requirements of KS’ Masters dissertation, supervised by JG and KCL. JG, KL, KS, CSE and FB conceived of the original idea. KS, JG and KL contributed to the design of the study. KS led, and KCL and JG contributed to the establishment of the search strategy and method of analysis. The original article reviewing and data analysis were led by KS, with contribution from KCL and JG. The updated article review and data analysis were led by JG, with contribution from KCL and KS. All authors (KS, KCL, CSE, FB and JG) have made contributions to the drafting and revising of the article, and have approved the final version.

