## Supplementary files for "Scoping review of online postal sexually transmitted infection services: access, usage and clinical outcomes"

### Supplementary File

Supplemental File 1: Search Strategy (where applicable limited to publications in English language, 2010-2020).

| Database | Medical Subject Headings | Keywords |
| --- | --- | --- |
| Medline | exp Sexually Transmitted Diseases, exp Sexual Health, exp Telemedicine, exp Internet, exp Health Services Accessibility, exp Healthcare Disparities, exp Socioeconomic Factors, exp Minority Groups, exp Social Marginalization, exp United Kingdom  NOT exp Africa, exp americas, exp antarctic regions, exp arctic regions, exp asia, exp oceania | "sexually transmitted disease*", "sexually transmitted infection*", sti, stis, std, stds, venereal, "sexual health*", chlamydia, gonorrhoea, gonorrhoea, syphilis, "herpes genitalis", hiv, "human immunodeficiency virus*", "acquired immunodeficiency syndrome*", aids, "acute retroviral syndrome*", ars, "hepatitis B", "hepatitis C" telemedicine*, telehealth*, "mobile health*", mhealth*, ehealth*, internet, "digital service*", "digital intervention*", "digital technolog*", "self sampl*", "self test*", "home test*", "test* kit*", "self collect*", "home base*", "web-base*", "self-swab*", "home swab*", "mobile technolog*", "mobile application*", app, apps, "social medi*", "cell phone*", cellphone*, "mobile phone*", "mobile telephone*", "cellular phone*”, smartphone*, "smart phone*", "mobile device*", "text messag*", texting, texted, sms, mms, "multimedia messag*", "short messag*", "computers, handheld", "personal digital assistant", email*, "e-mail*", online, "digital health*", access*, disparit*, barrier*, availab*,  inaccess*, unavailab*, socioeconomic*, minorit*, inequalit*, equalit*, inequit*, "equit", "marginaliz*", "marginalis*", "convenien*", "inconvenien*", "hard to reach", "national health service*", nhs* , gb, "g.b.", britain*, british*, uk, "u.k.", "united kingdom*", england*, “english”, "northern ireland*", "northern irish*", scotland*, scottish*, wales, “south wales”, welsh*  NOT “british columbia”, “new england”, “new south wales”, ((published or publication* or translat* or written or language* or speak* or literature or citation*) adj5 english) |
| EMBASE | exp sexually transmitted disease, exp sexual health, exp telemedicine, exp internet, exp health care access, exp health care disparity, exp socioeconomic, exp minority group, exp social exclusion, exp United Kingdom  NOT exp africa, exp americas, exp antarctic regions, exp arctic regions, exp asia, exp oceania | "sexually transmitted disease*", "sexually transmitted infection*", sti, stis, std, stds, venereal, "sexual health*", chlamydia, gonorrhoea, gonorrhoea, syphilis, "herpes genitalis", hiv, "human immunodeficiency virus*", "acquired immunodeficiency syndrome*", aids, "acute retroviral syndrome*", ars, "hepatitis B", "hepatitis C" telemedicine*, telehealth*, "mobile health*", mhealth*, ehealth*, internet, "digital service*", "digital intervention*", "digital technolog*", "self sampl*", "self test*", "home test*", "test* kit*", "self collect*", "home base*", "web-base*", "self-swab*", "home swab*", "mobile technolog*", "mobile application*", app, apps, "social medi*", "cell phone*", cellphone*, "mobile phone*", "mobile telephone*", "cellular phone*”, smartphone*, "smart phone*", "mobile device*", "text messag*", texting, texted, sms, mms, "multimedia messag*", "short messag*", "computers, handheld", "personal digital assistant", email*, "e-mail*", online, "digital health*", access*, disparit*, barrier*, availab*,  inaccess*, unavailab*, socioeconomic*, minorit*, inequalit*, equalit*, inequit*, "equit", "marginaliz*", "marginalis*", "convenien*", "inconvenien*", "hard to reach", "national health service*", nhs* , gb, "g.b.", britain*, british*, uk, "u.k.", "united kingdom*", england*, “english”, "northern ireland*", "northern irish*", scotland*, scottish*, wales, “south wales”, welsh*,  NOT “british columbia”, “new england”, “new south wales”, ((published or publication* or translat* or written or language* or speak* or literature or citation*) adj5 english) |
| PsycInfo  HMIC | exp sexually transmitted diseases, exp sexual health, exp telemedicine, exp internet, exp health care access, exp health disparities, exp socioeconomic status, exp minority groups | "sexually transmitted disease*", "sexually transmitted infection*", sti, stis, std, stds, venereal, "sexual health*", chlamydia, gonorrhoea, gonorrhoea, syphilis, "herpes genitalis", hiv, "human immunodeficiency virus*", "acquired immunodeficiency syndrome*", aids, "acute retroviral syndrome*", ars, "hepatitis B", "hepatitis C" telemedicine*, telehealth*, "mobile health*", mhealth*, ehealth*, internet, "digital service*", "digital intervention*", "digital technolog*", "self sampl*", "self test*", "home test*", "test* kit*", "self collect*", "home base*", "web-base*", "self-swab*", "home swab*", "mobile technolog*", "mobile application*", app, apps, "social medi*", "cell phone*", cellphone*, "mobile phone*", "mobile telephone*", "cellular phone*”, smartphone*, "smart phone*", "mobile device*", "text messag*", texting, texted, sms, mms, "multimedia messag*", "short messag*", "computers, handheld", "personal digital assistant", email*, "e-mail*", online, "digital health*", access*, disparit*, barrier*, availab*,  inaccess*, unavailab*, socioeconomic*, minorit*, inequalit*, equalit*, inequit*, "equit", "marginaliz*", "marginalis*", "convenien*", "inconvenien*", "hard to reach", "national health service*", nhs* , gb, "g.b.", britain*, british*, uk, "u.k.", "united kingdom*", england*, "northern ireland*", "northern irish*", scotland*, scottish*, wales, “south wales”, welsh*  NOT “british columbia”, “new england”, “new south wales” |
| CINAHL Plus | (MH "Sexually Transmitted Diseases+"), (MM "Sexual Health"), (MH "Telemedicine+"), (MH "Internet+"), (MH "Health Services Accessibility+"), (MM "Healthcare Disparities"), (MH "Socioeconomic Factors+"), (MM "Minority Groups"), (MH “United Kingdom”) | "sexually transmitted disease*", "sexually transmitted infection*", sti, stis, std, stds, venereal, "sexual health*", chlamydia, gonorrhoea, gonorrhoea, syphilis, "herpes genitalis", hiv, "human immunodeficiency virus*", "acquired immunodeficiency syndrome*", aids, "acute retroviral syndrome*", ars, "hepatitis B", "hepatitis C" telemedicine*, telehealth*, "mobile health*", mhealth*, ehealth*, internet, "digital service*", "digital intervention*", "digital technolog*", "self sampl*", "self test*", "home test*", "test* kit*", "self collect*", "home base*", "web-base*", "self-swab*", "home swab*", "mobile technolog*", "mobile application*", app, apps, "social medi*", "cell phone*", cellphone*, "mobile phone*", "mobile telephone*", "cellular phone*”, smartphone*, "smart phone*", "mobile device*", "text messag*", texting, texted, sms, mms, "multimedia messag*", "short messag*", "computers, handheld", "personal digital assistant", email*, "e-mail*", online, "digital health*", access*, disparit*, barrier*, availab*,  inaccess*, unavailab*, socioeconomic*, minorit*, inequalit*, equalit*, inequit*, "equit", "marginaliz*", "marginalis*", "convenien*", "inconvenien*", "hard to reach", "national health service*", nhs* , gb, "g.b.", britain*, british*, uk, "u.k.", "united kingdom*", england*, "northern ireland*", "northern irish*", scotland*, scottish*, wales, “south wales”, welsh*  NOT “british columbia”, “new england”, “new south wales” |
| Scopus  Open Grey  Web of Science | n/a | "sexually transmitted disease*", "sexually transmitted infection*", sti, stis, std, stds, venereal, "sexual health*", chlamydia, gonorrhoea, gonorrhoea, syphilis, "herpes genitalis", hiv, "human immunodeficiency virus*", "acquired immunodeficiency syndrome*", aids, "acute retroviral syndrome*", ars, "hepatitis B", "hepatitis C" telemedicine*, telehealth*, "mobile health*", mhealth*, ehealth*, internet, "digital service*", "digital intervention*", "digital technolog*", "self sampl*", "self test*", "home test*", "test* kit*", "self collect*", "home base*", "web-base*", "self-swab*", "home swab*", "mobile technolog*", "mobile application*", app, apps, "social medi*", "cell phone*", cellphone*, "mobile phone*", "mobile telephone*", "cellular phone*”, smartphone*, "smart phone*", "mobile device*", "text messag*", texting, texted, sms, mms, "multimedia messag*", "short messag*", "computers, handheld", "personal digital assistant", email*, "e-mail*", online, "digital health*", access*, disparit*, barrier*, availab*,  inaccess*, unavailab*, socioeconomic*, minorit*, inequalit*, equalit*, inequit*, "equit", "marginaliz*", "marginalis*", "convenien*", "inconvenien*", "hard to reach", "national health service*", nhs* , gb, "g.b.", britain*, british*, uk, "u.k.", "united kingdom*", england*, "northern ireland*", "northern irish*", scotland*, scottish*, wales, “south wales”, welsh*  NOT “british columbia”, “new england”, “new south wales” |
| Ethos | n/a | 1. “sexually transmitted infection” AND test 2. “sexually transmitted infection” AND online 3. “sexually transmitted infection” AND access 4. “sexually transmitted infection” AND UK 5. “sexually transmitted disease” AND test 6. “sexually transmitted disease” AND online 7. “sexually transmitted disease” AND access 8. “sexually transmitted disease” AND UK |

Supplemental File 2: Systematic Review Data Extraction Form (blank)

Systematic Review Data Extraction Form

| Record no. |
| --- |
| Article citation |
| Type: |
| Completed by: |

Summary Notes

Study Description

| Study aims/research questions |
| --- |
| Rationale (why did the study author believe the study could benefit the study population in this context?) |
| Participants (number and description) |
| Setting (geographic location, institutional setting, other place/space-related features) |
| Summary of intervention |

Data Collection

| What types of data are collected/managed/shared? |
| --- |
| How is this data collected/managed/shared? (describe the tool, if there is one) |
| Data collection and sharing context (who is collecting data and who is it being shared with, how?) |

Study Design

| Description of study design |
| --- |
| Theoretical framework for development and/or evaluation (describe, if there is one) |
| Eligibility (inclusion/exclusion criteria) |
| Control/comparison group |
| Analyses conducted |
| Outcome measures |

Study Findings (add more rows to elaborate on findings as needed)

| Key descriptive statistics |
| --- |
| Clinical outcomes |
| Other self-reported outcomes |
| Acceptability |

Assessment & Impacts

| Strengths and Limitations (as noted by authors) |
| --- |
| MMAT Assessment Results |
| Key impacts and lessons learned |

Key Quotes (for thematic analysis, add more rows as needed)

| Notes | Quote |
| --- | --- |

| **Supplemental File 3: Demographics of Study Participants** | | | | | | | |
| --- | --- | --- | --- | --- | --- | --- | --- |
| **Service** | Author | Study population & number of participants | Gender | Age | Ethnicity | Sexual orientation | Index of deprivation |
| **eSexual Health Clinic (eSHC)** | Aicken et al. 2018 (37) | Telephone interviews with patients who had used the eSHC  N=36 | Female - 20  Male - 16 | 18-24yrs – 18  25-35yrs - 18 | Asian - 3  Black - 7  Mixed - 4  White - 22 | Heterosexual – 34  Unstated - 2 | Unstated |
|  | Estcourt et al. 2017 (38) | Recruited from GUM clinic - 116 | Female - 74  Male - 42 | Median 25yrs | White British – 37  White other – 29  Black – 17  Asian, mixed or other – 21  Unstated - 12 | Same-sex partner in last 6 months - 1 | Unstated |
|  |  | Recruited from NCSP Checkurself - 105 | Female - 60  Male - 45 | Median 22yrs | White British – 67  White other – 5  Black – 12  Asian, mixed or other – 10  Unstated - 11 | Same-sex partner in last 6 months – 3 | Unstated |
|  | Gibbs et al. 2018 (39) | 1) Patients completing telephone survey - 152  2) Interviews (see Aicken et al. 2018)  3) Online survey - 331 | Unstated | Unstated | Unstated | Unstated | Unstated |
| **Freetest.me** | Dolan et al. 2014 (40) | 2988 | Male: 33.7  Female:66.3 | (Mean)  16-19yrs – 30.7  20-24yrs – 66.4  Unstated – 2.9 | (Mean)  White – 86.5  Black – 1.4  Asian – 1.3  Other – 0.3  Mixed – 3.1  Unstated – 7.3 | Unstated | (Mean)  IMD Score (SD) – 20.2 (14.13) |
| **Letstalkaboutit** | Gasmelsid et al. 2021 (34) | Before online testing  2847 | Female 1561  Male 1285 | <25 1949  25+ 898 | Black and Minority Ethnic: 209  White: 2099 | Non-heterosexual 120  Heterosexual 1336 | Unstated, but reported as no difference between groups |
|  |  | After online testing  2066 clinic | Female 1007  Male 891 | <25 1334  25+ 732 | Black and Minority Ethnic: 227  White: 1540 | Non-heterosexual 51  Heterosexual 875 |  |
|  |  | After online testing  775 online | Female 437  Male 210 | <25 520  25+ 255 | Black and Minority Ethnic: 33  White: 366 | Non-heterosexaul 28  Heterosexual 213 |  |
| **NCSP** | Woodhall et al. 2012 (5) | Internet test kits requested (number of participants unstated) – 59719 | Female: 38268 | 15yrs – 442  16-19yrs – 13099  20-24yrs – 24727 | White – 32604  Black – 796  Asian – 747  Chinese – 74  Other – 91  Mixed – 1032  Unstated – 2924 | Unstated | IMD quintile  1 (most deprived) – 6662  2 – 7965  3 – 7987  4 – 7341  5 – 7019  Unstated – 1294 |
|  |  |  | Male: 21451 | 15yrs – 237  16-19yrs – 6659  20-24yrs – 14555 | White – 17925  Black – 499  Asian – 519  Chinese – 24  Other – 59  Mixed – 569  Unstated – 1855 | Unstated | IMD quintile  1 (most deprived) – 3733  2 – 4276  3 – 4353  4 – 4093  5 – 4164  Unstated – 832 |
|  |  | GP tests (number of participants unstated) – 148619 | Female: 109,187 | 15yrs – 3322  16-19yrs – 41698  20-24yrs – 64167 | White – 67934  Black – 5851  Asian – 5464  Chinese – 721  Other – 540  Mixed – 2670  Unstated – 26007 | Unstated | IMD quintile  1 (most deprived) – 26319  2 – 24489  3 – 18318  4 – 14620  5 – 15565  Unstated – 9876 |
|  |  |  | Male: 39432 | 15yrs – 901  16-19yrs – 14283  20-24yrs – 24248 | White – 20976  Black – 2784  Asian – 4744  Chinese – 335  Other – 298  Mixed – 950  Unstated – 9345 | Unstated | IMD quintile  1 (most deprived) – 10837  2 – 9525  3 – 6186  4 – 4571  5 – 4495  Unstated – 3818 |
|  |  | SRH tests (number of participants unstated) – 202028 | Female: 156,432 | 15yrs – 13463  16-19yrs – 82264  20-24yrs –60705 | White – 97153  Black – 11886  Asian – 4127  Chinese – 510  Other – 733  Mixed – 6139  Unstated – 35884 | Unstated | IMD quintile  1 (most deprived) – 50020  2 – 35086  3 – 24394  4 – 19514  5 – 15441  Unstated – 11977 |
|  |  |  | Male: 45596 | 15yrs – 3215  16-19yrs – 24866  20-24yrs – 17515 | White – 26261  Black – 4002  Asian – 1488  Chinese – 75  Other – 194  Mixed – 1703  Unstated – 11783 | Unstated | IMD quintile  1 (most deprived) – 15331  2 – 10053  3 – 6395  4 – 5523  5 – 3952  Unstated – 4342 |
| **North East Essex PCT** | Bracebridge et al. 2012 (41) | People offered screening - 29917 | Female: 14773  Male: 15136  Unstated: 8 | 17-18yrs – 3773  19yrs – 4512  20yrs – 4532  21yrs – 4643  22yrs – 4220  23yrs – 4099  24-25yrs – 4128  Unstated – 10 | Unstated | Unstated | IMD quintile  1 (least deprived)* – 5857  2 – 6048  3 – 6004  4 – 5992  5 (most deprived)* – 6002  Unstated – 14 |
|  |  | People who completed screening - 3431 | Female: 1951  Male: 1480 | 17-18yrs – 466  19yrs – 564  20yrs – 569  21yrs – 466  22yrs – 444  23yrs – 434  24-25yrs – 486  Unstated – 2 | White – 2967  Other – 216  Unstated – 248 | Unstated | IMD quintile  1 (least deprived)* – 389  2 – 802  3 – 872  4 – 687  5 (most deprived)* – 676  Unstated – 5 |
|  |  | People who tested positive for chlamydia - 152 | Female: 85  Male: 67 | 17-18yrs – 18  19yrs – 21  20yrs – 20  21yrs – 20  22yrs – 32  23yrs – 21  24-25yrs – 20 | White – 135  Other – 6  Unstated – 11 | Unstated | IMD quintile  1 (least deprived)* – 11  2 – 39  3 – 26  4 – 42  5 (most deprived)* – 34 |
| **Saving Lives** | Page et al. 2019 (42) | Users of Mini-tube - 275 | Female – 166  Male – 106  Trans – 2  Unspecified – 0 | Median 26, IQR 22, 31  Mean 28, 95% CI 27, 29 | Any other mixed background – 2  Any other white background – 7  Bangladeshi – 1  Black African – 0  Black Caribbean - 0  Chinese - 0  Indian – 1  Unknown/Not specified – 3  White & Asian – 4  White and black African – 2  White and black Caribbean - 3  White British - 242  White Irish - 10 | Heterosexual Male – 86  Heterosexual Female – 152  MSM - 20  WSW - 16  Unknown/not spec - 1 | Unstated |
|  |  | Users of dried blood spot - 275 | Female – 94  Male – 181  Trans – 0  Unspecified – 0 | Median 25, IQR 22, 30  Mean 28, 95% CI 27,29 | Any other mixed background – 2  Any other white background – 5  Bangladeshi – 0  Black African – 1  Black Caribbean - 1  Chinese - 2  Indian – 0  Unknown/Not specified – 1  White & Asian – 3  White and black African – 0  White and black Caribbean - 1  White British - 253  White Irish - 6 | Heterosexual Male – 66  Heterosexual Female – 167  MSM - 28  WSW - 14  Unknown/not spec - 0 | Unstated |
| Sexual Health London | Day et al. 2020  (43) | 250 reported recent sexual assault on online triage form | Female: 135  Male: 114  Trans or non-binary: 1 | Mean 26  Range 18-55 | African – 28  Asian – 24  Caribbean – 23  Other – 64  White – 111 | Heterosexual men – 75  Heterosexual women – 105  MSM – 30  WSW – 2  Bisexual men – 9  Bisexual women – 28  Undetermined - 1 | Unstated |
|  | Day et al. 2021 (44) | 118,825 registrants identifying as cisgender, | Cisgender F:66955  Cisgender M: 51870 | Median age 27  Range 15-82 | African – 5537  Asian – 5781  Caribbean – 6417  Other – 17,482  White – 39082 | Unstated | Unstated |
|  |  | 504 registrants identifying as transgender, non-binary or ‘other’ | Trans men 76  Trans women 78  Non-binary/gender-fluid 17  Other 33 | Median age 27 Range 16-82 | African – 14  Asian – 21  Caribbean – 12  Other - 100  White - 145 | Unstated | Unstated |
|  | Day et al. 2020 (45) | Call back group  193 | Female: 133  Male: 57  Trans/non-binary: 3 | 16: 45  17: 148 | White: 81  Caribbean: 45  African: 26  Asian: 14  Other ethnicity: 27 | Heterosexual - 157  Homosexual – 15  Bisexual - 21 | Unstated |
|  |  | Non-call back group  261 | Female: 179  Males: 90  Trans/non-binary: 1 | 16: 74  17: 187 | White: 87  Caribbean: 90  African: 27  Asian: 15  Other ethnicity: 42 | Heterosexual - 227  Homosexual – 9  Bisexual - 25 | Unstated |
|  | Day et al. 2021 (46) | 34 confirmed new HIV results from 144 people with reactive HIV results. | Female: 2  Male: 32 | Median 28  Range 21-50 | White British or White Other: 20  Black, Asian and Minority Ethnic groups: 4  Other: 10 | Gay and bisexual MSM – 30  Heterosexual men – 2  Heterosexual women - 2 | Unstated |
| **SH:24** | Barnard et al. 2018 (32) | 5747 (3198 clinic, 2549 online) | Female: 3258  Male: 2489 | 16-20yrs – 358  20-25yrs – 1516  25-30yrs – 1798  30-35yrs – 895  35+yrs - 1180 | White British – 2233  White other – 829  Black African – 560  Black Caribbean – 480  Black other – 496  Mixed white black African or Caribbean’ – 225  South Asian – 90  Other – 600  Unstated - 229 | Heterosexual – 4731  Homosexual – 651  Bisexual – 165  Unstated - 200 | IMD quintile  1 (most deprived) – 2115  2 – 2394  3 – 974  4 – 219  5 (least deprived) – 29 |
|  | Barnard 2020 (Chapter 6) (47) | 20 | Female: 7  Male: 12  Trans: 1 | 16-19yrs – 1  20-24yrs – 10  25-30yrs - 9 | White – 8  Mixed – 3  Asian – 2  Black – 6  Other - 1 | Heterosexual – 13  Homosexual – 5  Bisexual - 2 | Unstated |
|  | Syred et al. 2019 (48) | Pre-intervention - 6253 | Female: 4030  Male: 2223 | 16-24yrs – 3351 | Black and Minority Ethnic – 642 | Men who have sex with Men (MSM) – 367 | IMD quintile  1 (most deprived) – 580  2 – 1364  3 – 1399  4 – 1614  5 (least deprived) - 1296 |
|  |  | Post- intervention - 7772 | Female: 4968  Male: 2804 | 16-24yrs – 4120 | Black and Minority Ethnic – 878 | MSM – 530 | IMD quintile  1 (most deprived) – 794  2 – 1705  3 – 1847  4 – 1886  5 (least deprived) - 1538 |
|  | Turner et al. 2018 (20) | Camberwell Sexual Health Clinic – 4172 | Female: 1340  Male: 921 | 16-19yrs – 201  20-24yrs – 472  25-29yrs – 508  30-34yrs – 365  35+yrs – 715 | White – 825  Mixed – 178  Asian – 59  Black or Black British – 939  Other – 208  Unstated - 52 | MSM – 251 | Unstated |
|  |  | Spontaneous SH:24 – 5632 | Female: 2746  Male: 1516 | 16-19yrs – 194  20-24yrs – 1282  25-29yrs – 1605  30-34yrs – 650  35+yrs – 531 | White – 2850  Mixed – 353  Asian – 107  Black or Black British – 768  Other – 105  Unstated - 79 | MSM – 505 | Unstated |
|  |  | Triage – 1266 | Female: 416  Male: 474 | 16-19yrs – 55  20-24yrs – 205  25-29yrs – 262  30-34yrs – 146  35+yrs – 222 | White – 461  Mixed – 93  Asian – 21  Black or Black British – 270  Other – 32  Unstated - 13 | MSM – 94 | Unstated |
|  | Turner et al. 2019 (33) | Time period 1 – 43491 clinic visits  Time period 2 – 51191 clinic visits, 11768 online orders | Unstated | Unstated | Unstated | Unstated | Unstated |
|  | Wilson et al. 2017 (49) | Intervention group - 1031 | Female: 604  Male: 424  Trans: 3 | 16-19yrs – 206  20-24yrs – 440  25-30yrs – 385 | White – 779  Black/African/Caribbean/black British – 81  Asian/Asian British – 70  Mixed – 89  Other – 12 | MSM – 129  Other – 890  Unstated – 12 | Unstated |
|  |  | Control group - 1032 | Female: 609  Male: 422  Trans: 1 | 16-19yrs – 220  20-24yrs – 432  25-30yrs – 380 | White – 749  Black/African/Caribbean/black British – 110  Asian/Asian British – 57  Mixed – 99  Other – 17 | MSM – 133  Other – 888  Unstated – 11 | Unstated |
|  | Wilson et al. 2019 (50) | Intervention group - 244 | Female: 130  Male: 113  Trans: 1 | 16-19yrs – 96  20-24yrs – 96  25-30yrs – 52 | White/WB – 176  Black/black British – 18  Asian/Asian British – 33  Mixed – 14  Other – 3 | MSM – 14  Other – 224  Unstated – 6 | Unstated |
|  |  | Control group - 284 | Female: 142  Male: 141  Trans: 1 | 16-19yrs – 118  20-24yrs – 110  25-30yrs – 56 | White/WB – 176  Black/black British – 25  Asian/Asian British – 34  Mixed – 26  Other – 5 | MSM – 21  Other – 258  Unstated – 5 | Unstated |
| **TakeATestUK.com** | Page et al. 2021 (51) | Mini tube - 1515 | Female:1051  Male:460  Trans:0  Other:4 | Mean (95% CI): 27.4 (27.1-27.8)  Median (IQR): 26 (22-31) | Any other Asian – 2  Any other Black – 6  Any other mixed – 11  Any other white – 41  Bangladeshi – 6  Black African – 25  Black Caribbean – 81  Chinese – 1  Indian – 5  Pakistani – 13  Unknown/not specified – 26  White and Asian – 17  White and Black African – 2  White and Black Caribbean – 79  White British – 1147  White Irish - 7 | Heterosexual man – 353  Heterosexual woman – 977  Bisexual man – 20  Bisexual woman – 18  MSM exclusive – 87  Women who have sex with women (WSW) exclusive – 56  Heterosexual trans woman - 0  Unknown/not specified – 4 | Unstated |
|  |  | Dried blood spot - 4155 | Female: 2788  Male: 1357  Trans: 2  Other: 8 | Mean (95% CI): 27.3 (27.1-27.5)  Median (IQR): 26 (2-31) | Any other Asian – 2  Any other Black – 6  Any other mixed – 11  Any other white – 41  Bangladeshi – 6  Black African – 25  Black Caribbean – 81  Chinese – 1  Indian – 5  Pakistani – 13  Unknown/not specified – 26  White and Asian – 17  White and Black African – 2  White and Black Caribbean – 79  White British – 1147  White Irish - 7 | Heterosexual man – 1012  Heterosexual woman – 2617  Bisexual man – 67  Bisexual woman – 95  MSM exclusive 278  WSW exclusive – 76  Heterosexual trans woman - 2  Unknown/not specified – 8 | Unstated |
| **Umbrella** | Banerjee et al. 2018 (35) | Patients who requested home-based kits – 9258 | Female: 5986  Male: 3258  Trans: 14 | 16-24yrs – 6033  25+yrs – 3225 | White – 6648  Black/black British – 892  Asian/Asian British – 558  Other – 920  Unstated – 240 | Heterosexual male – 2606  Heterosexual female – 5986  MSM – 652  Trans – 14 | Unstated |
|  |  | Patients who returned home-based kits - 4475 | Femae: 3104  Male: 1367  Trans: 4 | 16-24yrs – 2868  25+yrs – 1607 | White – 3375  Black/black British – 351  Asian/Asian British – 184  Other – 394  Unstated – 171 | Heterosexual male – 1039  Heterosexual female – 3104  MSM – 328  Trans – 4 | Unstated |
|  |  | Patients tested in clinic - 19193 | Female: 10861  Male: 8306  Trans: 26 | 16-24yrs – 9654  25+ yrs – 9539 | White – 7996  Black/black British – 4026  Asian/Asian British – 2167  Other – 2160  Unstated – 2844 | Heterosexual male – 6602  Heterosexual female – 10662  MSM – 1675  Trans – 24  Bisexual – 199  WSW – 31 | Unstated |
|  | Banerjee et al. 2020 (36) | Patients who requested home-based testing kit – 16,611 | Female: 10686  Male: 5889  Trans: 36 | 16-24yrs – 8819  ≥25yrs – 7792 | White – 11,519  Black – 1692  Asian – 1148  Other – 1770  Not stated - 482 | Heterosexual (M) – 5746  Heterosexual (F) – 10,667  Trans – 36  MSM – 138  Bisexual – 18  WSW - 6 | Unstated |
|  |  | Patients who returned home-based testing kits with sufficient quality of blood for testing –  9033 | Female: 6004  Male: 3018  Trans: 11 | 16-24yrs – 4623  ≥25 years - 4419 | White – 6588  Black – 803  Asian – 473  Other – 971  Not stated - 198 | Heterosexual (M) – 2875  Heterosexual (F) – 5985  Heterosexual trans – 11  MSM – 138  Bisexual – 18  WSW - 6 | Unstated |
|  |  | Patients who had serological testing in clinic | Female: 8236  Male: 8422  Trans: 37 | 16-24yrs – 6616  ≥ 25 - 10079 | White – 6331  Black – 3296  Asian – 1910  Other – 1796  Not stated - 3362 | Heterosexual (M) – 6617  Heterosexual (F) – 8093  Heterosexual trans – 37  MSM – 1788  Bisexual – 95  WSW - 65 | Unstated |
|  | Manavi et al. 2017 (52) | Test kits requested – 5310  (number of participants unstated) | Female kits: 3513  Male kits: 1787  Trans kits: 10 | Unstated | Unstated | Unstated | IMD rank  <5000 – 1855  5000-14999 – 2095  15000+ - 1321 |
| Table Key:  IMD: Index of Multiple Deprivation Unstated: unstated/refused/missing data SD: standard deviation  MSM: men who have sex with men WSW: women who have sex with women SRH: Sexual and Reproductive Health  **Bracebridge et al. 2012 have labelled IMD quintile 1 as least deprived and IMD quintile 5 as most deprived in their paper. This may be an error and makes this data difficult to interpret. The corresponding author has been contacted for clarification.* | | | | | | | |

| Supplemental File 4: Study Summaries and Key Findings | | | | | | | |
| --- | --- | --- | --- | --- | --- | --- | --- |
| Service Title | Authors (date) | Study type | Study aims | Description of study design | | Inclusion/ exclusion criteria | Key results |
| eSexual Health Clinic (eSHC) | Aicken et al. 2018 (37) | Qualitative interviews | To understand use and experience of the eSHC to inform future evaluation and refinement | - Telephone interviews with patients who had used the eSHC - Framework analysis carried out with codes developed from the sSHC | | Excluded: symptomatic or coinfected patients, aged <16yrs, unable to read English, no phone number provided | - Key themes included rapidity, protecting privacy, choice and seeking peace of mind - Subthemes included technology constraints, concerns with accessing results in public, simple and discreet treatment collection being positive and the trustworthiness conferred by the pathway being integrated within the NHS |
|  | Estcourt et al. 2017 (38) | Non randomised, exploratory proof of concept study | To assess the safety and feasibility of eSHC | - Chlamydia positive patients contacted 2 weeks after receiving their diagnosis - Outcomes included appropriate care management, time to treatment and proportion of partners treated - Chlamydia-negative users sent a brief acceptability survey | | Excluded: symptomatic or coinfected patients, already receiving treatment for undiagnosed chlamydia, <16yrs, unable to read English | - ~75% of users accessed the pathway of whom 60% managed solely online - ~25% users contacted the helpline - Most patients collected treatment from their allocated pharmacy - The day after receiving diagnosis 76% of GUM patients and 67% of NCSP Checkurself patients had collected treatment - 29% of GUM patients and 24% of NCSP Checkurself patients accessed online health information |
|  | Gibbs et al. 2018 (39) | Mixed methods evaluation | To evaluate the eSHC | - Data collected on eSHC webpage interactions and a survey to establish acceptability for chlamydia positive patients - Qualitative interviews with 36 users (see Aicken et al. 2018) - Survey of chlamydia-negative users - Acceptability analysed descriptively | | See Estcourt et al. 2017 and Aicken et al. 2018 | - 82-92% of patients accessed results within 5 days. Of these, 97% accessed their results on the day they received the text - For key findings from qualitative interviews, see Aicken et al. 2018 - 91% of chlamydia positive patients surveyed were pleased with the service, 66% of previous testers preferred the eSHC, 99% felt they received sufficient health information - Chlamydia negative users: 98% of first-time testers were pleased with the service, 90% of previous testers would use the eSHC again - 53% of those preferred the eSHC to alternative services |
| Freetest.me | Dolan et al. 2014 (40) | Randomised experiment | To explore the effect on chlamydia test return rates of non-cash financial incentives, and the influence of socioeconomic status | - Test requests randomly allocated to intervention (incentive) or control (no incentive) - Test kits sent identifiable bar codes for incentive - Logistic regression was conducted using postcodes to identify socioeconomic status | | Included: individuals using site during the study period | - Those requesting kits mostly female, white, displayed higher risk behaviours (e.g. 2+ partners in the last 12 months), less deprived, over 40% had completed a chlamydia test/over 30% had tested positive in the last year - Return rate was 71%, only small differences between incentives, none statistically significant - Individuals aged 15-19 and of lower socioeconomic position less likely to return tests |
| Letstalkaboutit | Gasmelsid et al. 2021 (34) | Retrospective service evaluation | To determine whether online screening is accessible by those patients most at need by comparing the demographics and number of asymptomatic chlamydial infections detected online and in clinic. | - Single service data - Comparison of the demographic characteristics and number of asymptomatic chlamydial infections detected via an online postal self-sampling service and in clinic - Two time periods: Time 1- pre-introduction of online postal self-sampling ((Sept 2014-March 2015); Time 2 and post-introduction of the online service (Sept 2017-March 2018) | | Included:  People testing positive for chlamydia in Solent NHS Trust services (clinic-based and online) between September 2017-March 2018 | - The demographic characteristics of individuals accessing services was similar in clinic and online services, and remained stable between Time 1 and Time 2. - The majority of patients diagnosed were <25 years old, of white ethnicity, heterosexual and women. - There were no differences in IMD before those diagnosed in Time 1 and Time 2 - There was a significantly higher proportion of service users who identified as gay, bisexual or other men who have sex with men in Time 2 compared to Time 1 - There was a significantly higher proportion of service users of Black, Asian and Minority ethnicity in Time 2 compared to Time 1 - Patients diagnosed in clinic were significantly more likely to wait more than a week for treatment than those diagnosed through online services. |
| NCSP | Woodhall et al. 2012(5) | Quantitative retrospective data analysis; website evaluation | To describe and evaluate access to the NCSP’s online chlamydia testing service | - Chlamydia testing data analysed to describe trends and proportion of internet tests, and describe online testing by area - Descriptive comparative analyses conducted of online vs face-to-face users - 90 websites evaluated on health promotion and clinical signposting information | | Included: had test codes for internet testing, aged 16-24 | - 5.3% of tests ordered online 2006-2010 - <0.5% in 2006 to a maximum of 7.1% in 2009 (varied by area, some <1% and others <40%) - Online users more likely men, aged 20-24, of white ethnicity and less deprived - Women more likely to have had a new sexual partner in the 3 months before testing, or +1 in the previous year, than face-to-face users - A high proportion of online tests resulted in positive chlamydia diagnoses - Internet testers spread out in levels of deprivation, whereas face-to-face testers were more deprived |
| North East Essex PCT | Bracebridge et al. 2012 (41) | Cross-sectional study | To quantify uptake and test-positivity rates, identify factors associated with screening and compare costs of the intervention with the NCSP | - Associations examined between personal characteristics and study outcomes: test uptake, service registration and test positivity - Demographic data obtained through test registration | | Included: aged 18-24, residing within the boundaries of NE Essex PCT | - 82% requested online screening - Screening uptake less likely among men, less deprived and over 20’s - Having 2+ partners in the previous year strongly associated to a positive diagnosis - 95.4% of chlamydia positive individuals and all notified partners requested postal treatment - Costs per screening test and positive diagnosis were 1.66 and 3.5 times more than the NCSP |
| Saving lives | Page et al. 2019 (42) | Observational study | To ascertain how DBS HIV kits compared with MT kits in this postal testing service. | - Single service dataset analysed to compare online requested MT and DBS HIV test kits - Analyses evaluated the online request, return and results of the two different bloods sampling techniques that were used sequentially by the service | | Included: All service users who ordered an HIV test kit during the study time period, and had consented to their anonymised data to be shared by a third- party organisation. | - Similar demographic characteristics of those accessing MT compared to those accessing DBS: 63% women, 90% white British, 86% heterosexual, median age 26) - No difference in return rates between MT and DBS. However statistically significant higher proportion of successful sample processing with DBS (98.8%) compared with MT (55.7%). - Higher proportion of false positives with MT (5.4%) *c.f.* DBS (0.0%) |
| Sexual Health London | Day et al. 2020 (43) | Retrospective service evaluation | To report the rate of recent sexual assault (SA) disclosure amongst users of SHL, and identify the outcomes of their call back discussions. | - Single service dataset - Service users reporting that they have been a victim of a recent sexual assault are contacted by the health advisor team - Outcomes included successful phone contact with patient, referral to Sexual Assault Referral Centre, Intervention by SHL team, test kit return, diagnosis of STI | | Included: All patients using SHL between 01/01/2020 and 18/02/2020 who triggered a call back for sexual assault | - 0.5% (242/45841) users triggered at least one call back for a SA - Majority of users were female (54.0%), heterosex­ual (72%), of white ethnicity (44.4%) and 80.4% had attended a sexual health clinic previously. - 79.3% (192/242) of call backs were successfully. - Of those that were contactable, 45% (87/192) of confirmed a recent SA and 52.6% (101/192) stated that they had made an error on the triage. 76.2% (77/101) of the latter were male. - 92.6% (224/242) kits were dispatched, and of these 73.7% (165/224) kits were returned and tested during the study period - For 90% (78/87) of those reporting a SA, no onward referral was made. |
|  | Day et al. 2021 (44) | Retrospective service evaluation | To assess the sexual health needs, sexual practices, STI/HIV positivity and satisfaction rates of trans and non-binary users of Sexual Health London | - Single service dataset - Demographic characteristics and outcomes of service users identifying as transgender, non-binary/gender fluid or ‘other’ (TNB) registering to use SHL. - Outcomes included: sexual practices, sexual/reproductive healthcare needs and prior SHC at­tendance, service outcomes, STI test results and satis­faction scores. | | Included: All people identifying as TNB when registering for SHL between 20^th^ April 2019 and 31^st^ December 2019 | - 0.42% (540/119329) of registrants identified as transgender, non-binary/gender fluid or ‘other’. - 463 kits were placed, and 355 kits were returned from 302 unique users. No difference in kit return rate compared to cisgender individuals - The odds of being of Black, Asian or Minority Ethnicity were 1.2 times higher compared with cisgender individuals. - The odds of returning a blood sample were 1.6 times higher compared with cisgender in­dividuals - TNB service users were significantly more likely to engage in sex work, and reported similar rates of chemsex, group sex and fisting to that seen with MSM. - 95% (50/51) of users would recommend SHL to friends/family - 85.4% (70/82) gave a 5/5 star service rating |
|  | Day et al. 2020 (45) | Retrospective service evaluation | To report the safeguarding concerns and outcomes of 16-17year olds accessing SHL | - Single service data - SHL uses questions adapted from the Spotting the Signs proforma - Demographic characteristics and outcomes of those 16-17 year olds whose responses to the questions triggered a phone call ((‘call back’ (CB)) from a health advisor, compared with those that didn’t - Outcomes included: type & number of safeguarding triggers, CB outcomes, safeguarding outcomes among CB cases, STI test kits ordered and returned, STI test results. | | Included: All 16 and 17 year olds who triggered a CB from a health advisor | - 42.5% (193/454) service users triggered one CB, and 7 triggered 2 when ordering a second kit (i.e. they were 200 CBs triggered) - The most common reasons for triggering a call back were related to drug and alcohol use (27%0. Partner’s age imbalance (18% and involvement with social or mental health services (8%_ - All users received at least one CB attempt, and 84.5% had a successful call back. - 37.9% had a trigger downgraded (mainly because they had misread, misinterpreted or teicked the question in error. 6.5% disclosed additional or more serious concerns - 35.5% were referred to or attend a sexual health clinic - 8.5% were referred to the child protection team, mostly because they were not contactable or became uncontactable. - 7% of cases involved a discussion/referral to social services. |
|  | Day et al. 2021 (46) | Retrospective service evaluation | To identify the characteristics and transfer to care rates of those who have a reactive HIV test result via SHL | - Single service data - Demographics and outcomes of those users with a reactive HIV test result | | Included:  People with reactive HIV test results between 8^th^ January 2018-31^st^ December 2019 | - 0.097% (144/148,257) had a reactive HIV result - 20.8% (30/144) were known to be living with HIV - 29.8% (34/114) were confirmed HIV positive - 59.6% (68/114) were found to be HIV negative (i.e. were false positives) - In 10.5% (12/114) the result was unknown - Of those confirmed as new HIV diagnoses, all service users transitioned to a HIV outpatient clinic. The majority were male (94.1%), men who have sex with men (88.2%), and of white ethnicity (58.8%). |
| SH:24 | Barnard et al. 2018 (32) | Cross-sectional study | To compare the characteristics of e-STI service users with clinic users, and OPSS kit returners with non-returners | - Service activity data collected from sexual health clinics in Lambeth and Southwark - Complete case analysis carried out using logistic regression | | Included: residents of Lambeth and Southwark  Excluded: activity codes outside the remit, testing by prisoners | - Online services most popular with users aged 20-30 years, women, white British ethnicity, homosexual or bisexual individuals, those who receive negative results, and are less deprived - Women who were ‘mixed white black African or Caribbean’ had lower odds of using online services compared to men in this group - Homosexual women were more likely to use online services than homosexual men - The most likely groups to return samples were >20 years (p<0.05) and white British |
|  | Barnard 2020 (Chapter 6) (47) | Qualitative interviews | To describe the experiences, barriers and facilitators of SH:24 in Lambeth and Southwark | - Stratified purposive sampling of consenting trial participants (Wilson et al. 2017) - Thematic analysis | | See Wilson et al. 2017 | - Key themes were trust, subjective norms, privacy, self-efficacy, convenience and perceived risk of infection - Subthemes included service reliability and confidentiality, comfort and control, concealing testing, improved trust and subjective norms over time, risk of infection, self-sampling and validity of results |
|  | Syred et al. 2019 (48) | Observational study | To describe user choice of OPSS orders and diagnoses in a ‘choose to test’ intervention | - An online ‘choose to test’ intervention was piloted in Essex, UK - Users given a personalized test package - Users could add or remove tests - Outcomes included test package edits, cost of tests and diagnoses | | n/a | - Slight increase in MSM and more deprived groups occurred after the intervention - 17.2% of MSM removed tests, 67.3% of BME users added a syphilis test, 59.8% of users in neither group added HIV and syphilis tests - Orders from women and BME groups most likely to be modified, orders from 16-24s, MSM and symptomatic users least likely to be modified - Number of positive chlamydia or gonorrhoea diagnoses did not significantly change - Where users are given ‘choice to test’, most will choose chlamydia, gonorrhoea, HIV and syphilis - Costs are saved through a reduction in HIV and syphilis tests ordered |
|  | Turner et al. 2018 (20) | Observational study | To investigate the effect of decision-making on resource allocation in a clinic after the introduction of an e-STI service in Lambeth and Southwark | - Demographic data, type of STI test and area of residence were collected before and after the intervention - Outcomes analysed included testing volume and complexity in clinic, and test positivity between pathways | | Included: residents of Lambeth and Southwark only  Excluded: codes lacking from clinic visit, prisoners, <16yrs or >100yrs | - Online testing increased the volume of testing, clinic visits were proportionally more complex - The greatest proportion of 16-19s tested in clinic - Women were most likely to use the online service but less likely to return tests, MSM were comparatively likely to use clinic or e-STI services - Most online or triaged users were asymptomatic - reflective of the encouragement to use clinics if displaying symptoms - More tests returned by spontaneous online users than triage patients (p=0.01) |
|  | Turner et al. 2019 (33) | Case study analysis and model generation | To establish cost-effectiveness of an OPSS service, and explore cost per diagnosis in different scenarios | - Records of demographics, sexual orientation and clinical information from clinic visitations were captured - Online service data collected - Case study analysis conducted on cost-effectiveness of testing prior and post e-STI service | | Excluded: codes lacking from clinic visit, prisoners, <16yrs or >100yrs | - Clinic testing rates remained stable after SH:24 was introduced, but online testing increased overall testing volume by 27% from 2014-16 - 37% of tests were ordered online by 2016 - Average test positivity rates higher in clinic than online - Average monthly diagnoses and annual cost of testing increased from 2014-16, but cost per test and per diagnosis decreased - Return rates impact cost if they are below 60%; this study observed an over 75% return rate |
|  | Wilson et al. 2017 (49) | Single-blind randomised control trial (RCT) | To assess the effectiveness of an OPSS service compared with face-to-face services | - Participants recruited from community settings, online and through clinics - Participants randomly allocated to intervention (text message with e-STI service link) or control (text message with local clinic information) - Staff were blinded to the allocation - Outcomes included test completion, time to testing or treatment, positive tests and acceptability | Included: aged 16-30, resident in Lambeth or Southwark, 1+ sexual partner in the 12 months prior, willing to complete a test, internet access  Excluded: unable to read English or provide consent | | - STI testing at 6 weeks higher in the intervention group (50% vs 26.6%, p<0.001) with no evidence of heterogeneity across population groups so could be targeted to higher-risk groups - Time to test shorter in the intervention group (28.8 days vs 36.5 days, p<0.001) - Proportion of diagnoses or individuals treated, and time to treatment were not statistically significant (patients required to get treatment in clinic) - 71% surveyed found the intervention acceptable |
|  | Wilson et al. 2019 (50) | Secondary data analysis of an e-STI RCT in Lambeth and Southwark | To examine the effect of an e-STI service on testing uptake on people who had never previously tested (never-testers) | - Data analysed on never-testers - Outcomes included testing at 6 weeks, time to test, positive results, test completion by service, and acceptability - Interactions examined between demographics | | See Wilson et al. 2017 | - Data available for 87% of the intervention and 79% of the control group - Return of STI test at 6 weeks was higher for the intervention group (~45% vs ~25%, p<0.001) - Intervention reduced time to test at 42 days - Face-to-face community recruitment increased the effectiveness of the intervention - 74.6% surveyed found the intervention acceptable - There were greater proportions of men, 16-19 year old’s, and Asian or Asian British ethnicities in the never-testers - Further barriers - around 54% did not test |
| TakeATestUK.com | Page et al. 2021 (51) | Observational study | To ascertain how DBS HIV and syphilis kits compared with MT kits in this postal testing service | - Single service dataset analysed to compare online requested MT and DBS HIV test kits - Analyses evaluated the online request, return and results of the two different bloods sampling techniques that were used sequentially by the service. - Secondary objectives were to describe the number of kit requests to obtain one successfully processed result, and the proportion of false-positive results for DBS and MT. | | Included: All service users who ordered an HIV/syphilis test kit during the study time period, and had consented to their anonymised data to be shared by a third- party organisation. | - Similar demographic characteristics of those accessing MT compared to those accessing DBS (majority female (69.4% vs 67.1), white British (75.7% vs 73.2%) heterosexual (87.8% vs 87.3%), median age 26 (IQR 22-31) - No difference in proportion of people who returned STI kit who also returned HIV/STS sample (88.2% for MT, 87% for DBS, p= 0.340). - Statistically significant higher proportion of successful sample processing with DBS (94.6%) compared with MT (54.4%), p=<0.001. - Higher proportion of MT samples were reactive for HIV compared with DBS (6.2% vs 1.1%, p=<0.001). However, higher proportion of false positives with MT than DBS (5.2% of all successfully processed MT samples *c.f.* 0.4% of DBS) - Lower proportion of STS samples were reactive (1.1% for MT samples *c.f*.0.7% of DBS), with 0.4% of successfully processed MT samples being false positive *c.f*. 0.0% for DBS.) |
| Umbrella | Banerjee et al. 2018 (35) | Retrospective service evaluation | To evaluate the rates of uptake and return of OPSS kits and compare patient demographics and clinical outcomes in home and clinic testers | - Retrospective data compared between home test users and clinic test users - Outcomes included time before kit return, time to treatment, positivity and treatment rates | | Included: patients who took home or clinic based STI tests in the study period, >16yrs, residing in Birmingham/Solihull | - A third of patients requested home tests, 48% returned kits - Home sampling was more popular among those aged 16-24, of white ethnicity, heterosexual female and asymptomatic (p<0.001) - Home sampling was less popular in black/black British and Asian/British Asian groups (p<0.001) - Positivity rates higher in clinic - Treatment rates lower in the home self-sampling group and time-to-treat longer than in clinic groups |
|  | Banerjee et al. 2020 (36) | Retrospective service evaluation | To evaluate the uptake, return rate and new diagnosis rates of home-base testing in comparison with clinic-based for HIV, STS, and hepatitis B | - Retrospective data compared between home test users and clinic test users - Outcomes included total number of patients in the two groups according to patient demographics, and presence of symptoms, number of patients returning home kit with sufficient quantity of blood compared to serological testing in clinci, overall retunr rate of home-based testing kits, number of patients with reactive results using home-based kits compared to clinic, outcomes of patients with reactive/equivcal tests in both groups. | | Included:  Patients using home-based testing kits or attending clinics in the Birmingham & Solihull area between July-December 2017 | - Home sampling was more popular amongs those aged 16-24, white, female patients (p<0.001. - Home sampling was less popular in Black and Asian groups (p<0.001) - Only 54% (9033/16,611) of home-based test kits were returned with sufficient quantity of blood for testing. - False positivity rate was significantly higher for HIV and Hepatitis B in the home based group. - 26/75 of HIV, 71/146 of STS, and 12/15 Hep B reactive results from home-based sampling were unconfirmed. - Only 1/75 of reactive HIV tests, 8/146 for STS, and 0/15 for Hepatitis B from home-based test kits were true-positives, new cases. |
|  | Manavi et al. 2017 (52) | Observational study | To establish which factors influence return of OPSS kits | - Retrospective data collected including demographic information, sexual history and symptoms - This data linked to the laboratory system to confirm which individuals returned requested kits | | Included: use of the service within the study period | - 58.4% of kits were returned (61.2% of women vs 53.1% of men, p<0.001, and 10% of transgender individuals). MSM had similar rates of kit return to women; heterosexual men were less likely to return kits - Those who returned tests were less deprived - Patients requesting home kits rather than pharmacy-collection were more likely to return them - Symptomatic patients less likely to return kits, potentially due to clinic attendance |

Supplemental File 5 – Mixed Methods Appraisal Tool (MMAT)

| Author (date) | S1: Are there clear research questions? | S2: Do the collected data allow to address the research questions? | 1.1. Is the qualitative approach appropriate to answer the research question? | 1.2. Are the qualitative data collection methods adequate to address the research question? | 1.3. Are the findings adequately derived from the data? | 1.4. Is the interpretation of results sufficiently substantiated by data? | 1.5. Is there coherence between qualitative data sources, collection, analysis and interpretation? | Comments |
| --- | --- | --- | --- | --- | --- | --- | --- | --- |
| Aicken et al. 2018 (37) | YES | YES | 1.1 YES | 1.2 YES | 1.3 YES | 1.4 YES | 1.5 YES |  |
| Barnard 2020  (Chapter 6) (47) | YES | YES | 1.1 YES | 1.2 YES | 1.3 YES | 1.4 YES | 1.5 YES |  |
|  | | | 2.1. Is randomization appropriately performed? | 2.2. Are the groups comparable at baseline? | 2.3. Are there complete outcome data? | 2.4. Are outcome assessors blinded to the intervention provided? | 2.5 Did the participants adhere to the assigned intervention? |  |
| Dolan 2014 (40) | YES | YES | 2.1 YES | 2.2 YES | 2.3 YES | 2.4 NO | 2.5 YES |  |
| Wilson 2017 (49) | YES | YES | 2.1 YES | 2.2 YES | 2.3 YES | 2.4 YES | 2.5 YES |  |
| Wilson 2019 (50) | YES | YES | 2.1 YES | 2.2 YES | 2.3 YES | 2.4 YES | 2.5 YES | This was secondary analysis of RCT data. The MMAT has been completed based on the criteria of the original RCT (Wilson et al. 2017) |
|  | | | 3.1. Are the participants representative of the target population? | 3.2. Are measurements appropriate regarding both the outcome and intervention (or exposure)? | 3.3. Are there complete outcome data? | 3.4. Are the confounders accounted for in the design and analysis? | 3.5. During the study period, is the intervention administered (or exposure occurred) as intended? |  |
| Banerjee 2018 (35) | YES | YES | 3.1 YES | 3.2 YES | 3.3 YES | 3.4 NO | 3.5 YES |  |
| Banerjee 2020 (36) | YES | YES | 3.1 YES | 3.2 YES | 3.3 YES | 3.4 NO | 3.5 YES |  |
| Barnard 2018 (32) | YES | YES | 3.1 YES | 3.2 YES | 3.3 YES | 3.4 YES | 3.5 YES |  |
| Bracebridge 2012 (41) | YES | YES | 3.1 YES | 3.2 YES | 3.3 YES | 3.4 YES | 3.5 YES |  |
| Day 2020 (43) | YES | YES | 3.1 YES | 3.2. YES | 3.3. NO | 3.4 NO | 3.5. YES |  |
| Day 2021 (44) | YES | YES | 3.1 CAN’T TELL | 3.2. YES | 3.3. NO | 3.4. NO | 3.5 YES |  |
| Day 2020 (45) | YES | YES | 3.1. YES | 3.2. YES | 3.3. NO | 3.4 NO | 3.5 YES |  |
| Day 2021 (46) | YES | YES | 3.1. YES | 3.2 YES | 3.3. NO | 3.4 NO | 3.5 YES |  |
| Estcourt 2017 (38) | YES | YES | 3.1 YES | 3.2 YES | 3.3 YES | 3.4 NO | 3.5 YES |  |
| Gasmelsid 2021 (34) | YES | NO | 3.1 NO | 3.2. NO | 3.3. NO | 3.4 NO | 3.5 YES |  |
| Page 2019 (42) | YES | NO | 3.1 YES | 3.2 YES | 3.3. YES | 3.4 NO | 3.5 YES |  |
| Page 2021 (51) | YES | NO | 3.1 YES | 3.2 YES | 3.3. YES | 3.4 NO | 3.5 YES |  |
| Syred 2019 (48) | YES | YES | 3.1 YES | 3.2 YES | 3.3 YES | 3.4 YES | 3.5 YES |  |
| Turner 2018 (20) | YES | YES | 3.1 YES | 3.2 YES | 3.3 YES | 3.4 YES | 3.5 YES |  |
| Turner 2019 (33) | YES | YES | 3.1 YES | 3.2 YES | 3.3 YES | 3.4 NO | 3.5 YES |  |
|  | | | 4.1. Is the sampling strategy relevant to address the research question? | 4.2. Is the sample representative of the target population? | 4.3. Are the measurements appropriate? | 4.4. Is the risk of nonresponse bias low? | 4.5. Is the statistical analysis appropriate to answer the research question? |  |
| Manavi 2017 (52) | YES | YES | 4.1 YES | 4.2 YES | 4.3 YES | 4.4 YES | 4.5 YES |  |
| Woodhall 2012 (5) | YES | YES | 4.1 YES | 4.2 YES | 4.3 YES | 4.4 YES | 4.5 YES |  |
|  | | | 5.1. Is there an adequate rationale for using a mixed methods design to address the research question? | 5.2. Are the different components of the study effectively integrated to answer the research question? | 5.3. Are the outputs of the integration of qualitative and quantitative components adequately interpreted? | 5.4. Are divergences and inconsistencies between quantitative and qualitative results adequately addressed? | 5.5. Do the different components of the study adhere to the quality criteria of each tradition of the methods involved? |  |
| Gibbs 2018 (39) | YES | YES | 5.1 YES | 5.2 YES | 5.3 YES | 5.4 YES | 5.5 YES |  |

| Supplemental File 6: Summaries of the ten online postal STI services | | | |
| --- | --- | --- | --- |
| Service | Setting | | Summary of service or intervention |
| eSHC  (37–39) | Greater London | - Individuals undergo STI testing at one of two included genitourinary (GUM) clinics or via an online postal self-sampling service provided by six NCSP areas in South London - All eligible patients receive an discreetly worded text message from the secure NHS SMS system stating that their results are available and can be viewed using an attached link for a password-protected online application - Patients log on with their date of birth and either their clinic or phone number - If the result is positive, patients are offered and may provide consent for their use of a remote, self-directed online chlamydia pathway. They complete an online consultation to provide routinely collected clinical and public health surveillance data, and they are provided with trusted links to access information about their condition. Patients can then nominate one of 30 participating pharmacies from which they can collect their treatment - If users receive a negative result, health promotion material is provided - If users opt out of the online care pathway, they redirected to traditional face-to-face care - A helpline staffed by research health advisors is accessible from 9:00am-5:00pm on weekdays - If users describe factors such as allergies, symptoms or drug use during their online consultation then they are directed to call the helpline whose staff will facilitate their access directly to face-to-face care - Treatment for sexual partners is recommended; sexual partners can be notified and linked to the record of their sexual partner | |
| Freetest.me  (40) | All areas of England except within the North East Strategic Health Authority | - Preventx Limited provide an online and text service called Freetest.me - Individuals request a postal kit online; samples are returned by post using the prepaid return box - Individuals choose to be informed that their results are available for access by either text or email. They can opt in to being telephoned if results are positive - Results can be viewed online via an online tracking system - A reminder text is sent if samples have not been returned within 18 days, and samples not returned within 30 days of the request date are deemed invalid | |
| Letstalkaboutit (34) | Hampshire | - OPSS service offered to residents of Hampshire, through Solent NHS Trust. - Introduced county-wide in 2015. - Local sexual health clinics sign post eligible service users to the Letstalkaboutit website. Service users request a kit online, the kit is posted to their home address, and they post the samples back to the laboratory. - Results via text or phone. | |
| NCSP  (5) | England | - The English National Chlamydia Screening Programme provides no-cost opportunistic testing for young people and easy access to treatment - Sexually active patients attending healthcare settings such as a general practice (GP), local pharmacies, and sexual health, abortion or reproductive health services are offered point-of-care tests - Patients can also access tests via the internet, and some may receive postal invitations to test | |
| North East Essex PCT  (41) | North East Essex | - Home self-sampling kits for chlamydia posted to all young people (18-24years) within the PCT boundary. Tests included uniquely numbered containers to collect urine samples, instructions, information about service registration, informative material on sexual health and a prepaid envelope for kit return - Individuals hoping to use the service were directed to sign up with their unique number through either a webpage or using a freephone number - Individuals were notified of available results by text or other means requested and could log into their account - If receiving a positive result, patients completed an online questionnaire and chose whether they would like to collect their treatment from a pharmacy or have it posted to them. A doctor remotely reviewed this information and prescribed treatment if appropriate. - If the doctor was concerned by information provided in the questionnaire, they would contact the patient directly to discuss treatment - If individuals did not have internet access they could use the freephone number to seek assistance - Sexual partners could receive notification from the service and could be linked to existing cases | |
| Saving lives (42) | North-West of England sexual health clinic | - OPSS service provided by a charity (Saving Lives), their partners and Public Health England Birmingham Laboratories - Change of how self-samples are collected for HIV testing within this service occurred in August 2017 - Before August 2017, blood was collected using finger-prick capillary blood sampling into a 500µL mini-tube. From August 2017, blood was collecting using finger prick capillary blood sampling onto specialised filter paper (DBS). The volume of bloods required for DBS is much smaller than for MT. | |
| Sexual Health London (43–46) | 29 boroughs of London | - SHL is a consortium led by Preventx Limited, which provides the online testing service; the data controller is City of London; and the clinical governance lies with Chelsea and Westminster NHS Foundation Trust - SHL was launched in 2018, and is available to residents of the participating London boroughs who are aged 16 years or older. - Service users register for an account online, complete an online consultation and, if eligible, are either posted the test kit or are able to collect it from a local sexual health clinic (in some areas). Self-taken samples are posted back to the Preventx laboratory for testing. - Service users with symptoms, or other needs that mean they are unsuitable for remote testing, are signposted to their local sexual health service. - Local sexual health clinics refer suitable attendees to SHL. - Access results via online portal (or are phone reactive HIV results) | |
| SH:24  (20,32,33,47–50) | Lambeth and Southwark, London; Essex | - Individuals complete a short form on the SH:24 website to order a free postal self-sampling kit for chlamydia, gonorrhoea, HIV and syphilis; test kits for men who have sex with men (MSM) include rectal and pharyngeal swabs. The kits include pictorial leaflets and links to the SH:24 website which has guidance and videos of how to collect blood samples - Individuals reporting symptoms are advised to visit clinics but can use SH:24 postal kits if they prefer - Individuals reporting complex needs such as mental health conditions are spoken to by phone and are referred to relevant services but can still use SH:24 postal kits if they prefer - Users return kits in a prepaid envelope and are sent text messages to indicate the progress of their order. SH:24 contact information is provided in these messages for individuals who would like to discuss any concerns. - If SH:24 has not received the postal test within 2 weeks, a text reminder is sent and tests are resent if requested - STI test results for chlamydia, gonorrhoea and syphilis are sent by text and if they are positive then patients are signposted to clinics; HIV-reactive test results are provided by telephone call. - If results are positive, sexual partners can be notified | |
| TakeATestUK (51) | Midlands based sexual health clinic | - OPSS service provided by a charity (Saving Lives), their partners and Public Health England Birmingham Laboratories - Change of how self-samples are collected for HIV/STS testing within this service occurred in August 2017 - From 6^th^ December 2016- 1^st^ November 2017, blood was collected using finger-prick capillary blood sampling into a 500µL mini-tube. From 3^rd^ November 2017, blood was collecting using finger prick capillary blood sampling onto specialised filter paper (DBS). The volume of bloods required for DBS is much smaller than for MT. | |
| Umbrella  (35,52) | Birmingham and Solihull | - Individuals self-register and fill out an online questionnaire. They are then issued a self-sampling kit, either to be posted to their chosen address or available from Umbrella pharmacies or clinics - Patients with symptoms are advised to attend a face-to-face clinic, but are able to order a test online if they prefer - Patients collect their samples and post the kit to a laboratory - Individuals are offered STI screening services if they choose to attend an Umbrella clinic - If patients receive positive test results, despite which method was used for screening, they are recalled for their treatment. They receive this invitation by SMS, followed by further telephone or written contact if consent was provided | |
